# Evaluation of Sex Hormones in Autoimmune Diseases in Pre- and Post-Menopausal Women

**DOI:** 10.64898/2026.08.19.26360795

**Authors:** Hari Krishnamurthy, Yuanyuan Yang, Qi Song, Karthik Krishna, Vasanth Jayaraman, Tianhao Wang, Kang Bei, John J. Rajasekaran

## Abstract

Autoimmune diseases have shown biased proportion in female population, existing clinical investigations of sex hormones in autoimmune populations have been relatively limited in terms of patient size and types of hormones investigated. In this study, we examined the relationship of sexual hormones and autoimmune antibodies in a large cohort of US women. This retrospective study sample included a total of 15319 female subjects’ medical information that were collected between December 2015 to May 2019 and tested in the Vibrant America Clinical Laboratory. The present serum sample was limited to female participants who had ever menstruated at the time of blood collection and completed the testing of the autoimmune antibodies and sex hormones. We focused on a total of 13 clinically significant autoantibodies including antinuclear antibody (ANA), 11 anti-extractable nuclear antigens (anti-ENAs), anti-cyclic citrullinated peptide 3 (anti-CCP3), and 11 female sex hormones. First, the prevalence of serological autoantibodies in a large set of adult female subjects divided by the menopause age was investigated. Next, the levels of sex hormones were compared in the seropositive autoimmune subjects and seronegative controls across the pre- and post-menopausal female groups. The presented study involving a large cohort of females showed no statistically different levels of sex hormones in seropositive autoimmune subjects and matched controls except for DHEA-s.

**Strengths and limitations of this study:**

- For the first time, a large cohort of female subjects (N=15319) and their levels of 13 autoantibodies and 11 sex hormones were evaluated.
- When possible, we concluded major literatures in related topics and compared with the data reported in this study.
- Data were derived from a single laboratory, and the study was retrospective, which could have caused bias.

## INTRODUCTION

Autoimmune disease is characterized by a disruption of immune tolerance of self-antigens and resulting in damage or dysfunction of tissues. A wide range of autoimmune diseases have been recognized and they can be either systemic or can affect specific organs or body systems.^1^ A gender bias has been observed in majority of the autoimmune diseases and the number of autoimmune diseases is strikingly greater in females than in males.^2,3^ In systemic lupus erythematosus (SLE) and Sjögren’s syndrome (SS), female sex appears to be the strongest risk factor with a female-to-male sex ratio of 9:1.^4,5,6^ In rheumatoid arthritis (RA), the gender bias is relatively modest but still has a female-to-male ratio of 3:1.^7,8^ Several possible reasons have been proposed but no mechanism has been established for this sex specific differences. In fact, clinicians have observed females generally have superior humoral and cell-mediated immunity and are more resistant to a variety of infections. One widely accepted hypothesis believes that the generally stronger immune system in female also implicates a mere response to a variety of complex situations such as loss of self-tolerance and aberrant responses to self-antigens. Other possibilities include x-chromosome immune-related genes, fetal micro chimerism, and differential hormonal and reproductive factors.

Relatively scant attention has been gathered to study the influence of sex hormones in autoimmune diseases meanwhile results from several animal and human studies were disparate.^9^ Hormones are associated with innate, adaptive, humoral and cell-mediated immune responses, and dysfunction of the immune system may lead to immune-mediated diseases including autoimmune disease. The occurrence of autoantibodies indicates a high possibility of autoimmune diseases though it alone is not diagnostic.^10^ The cause of autoantibody production is varied and not well understood. There has been a growing interest to study the relationship between the presence of autoantibodies and sex hormones. Among all sex hormones, estrogen has been most extensively studied in its possible modulation role for autoimmune diseases. Estradiol, as the most predominant form of estrogen in serum, was reported to affect the activation of B cells which increases autoantibody production^11,12^ and promote survival of autoreactive B cells.^13^ Estradiol has been traditionally considered to contribute to SLE pathogenesis and worsen disease activity in mice and humans;^14^ however, previous studies assessed serum estradiol levels in SLE subjects and majority of the studies showed no difference between patients and controls.^15^ To the contrary, androgens have been shown to have immunosuppressive effects on the immune responses by reducing B cell autoantibody responses.^16,17^ Although several studies reported a trend of decreased level of testosterone concentrations in SLE patients, most of these studies did not reach statistical significance.^15^ Moreover, the role of progesterone and leptin in autoimmunity have also been investigated.

In this study, we aim to explore sex hormone levels in two groups female subjects – a group of high-level autoantibody carriers (seropositive, n=4717) and a group of normal subjects whose autoantibody levels are in range (seronegative, n=10602). A panel of 14 autoantibodies including antinuclear antibody (ANA), anti-dsDNA, anti-cyclic citrullinated peptide (anti-CCP), and 10 anti-extractable nuclear antigens (anti-ENAs) were chosen due to their validated clinical utility in autoimmune diseases. Eleven sex hormones including estradiol, testosterone, sex hormone-binding globulin (SHBG), prolactin, progesterone, dehydroepiandrosterone sulfate (DHEA-S), luteinizing hormone (LH), parathyroid hormone, follicle-stimulating hormone (FSH), Human Insulin-like Growth Factor I (HIGF1), and cortisol were individually examined between two groups. The prevalence of various high-level autoantibodies in the seropositive female subjects was investigated. Furthermore, the levels of the hormones were compared across seropositive and seronegative autoimmune subjects. In the light of the large sample size (n=15319), certain approximations were able to be made to focus on the function of hormones. First, depending on women’s hormonal cycles, estradiol, progesterone, FSH, LH levels may fluctuate more dramatically but the fluctuation is expected to be mitigated by the large size of subjects. Second, 50-year-old is used as the menopause age to divide each group. Our study shows that even though sex hormones might be an important factor of autoimmune disease based on epidemiologic evidence, the correlation seems to be indirect and pathogenesis pathway needs systemic investigation.

## MATERIALS AND METHODS

### Serum Samples

This retrospective study samples included a total of 15319 female subjects’ medical information that was collected between December 2015 to May 2019 and tested in the Vibrant America Clinical Laboratory. The waiver of consent for In Vitro Diagnostic Device study using leftover human specimens that are not individually identifiable was approved by the Western Institutional Review Board (WIRB) (work order #1-1098539-1). The serum samples were limited to female participants (N=15319) who had ever menstruated at the time of blood collection and completed the testing of 13 autoimmune antibodies and 11 sex hormones as elaborated below. None of the samples or data were collected from repeated subjects.

### Autoimmune Antibody Panel

The ANA detection was performed with a solid phase bio-chip immunofluorescence assay, Vibrant™ ANA HEp-2 (Vibrant America, LLC, Santa Clara, CA, USA). A sample was considered ANA positive (ANA+) if any specific staining (homogeneous, centromere, speckled, nucleolar, peripheral) was observed to be greater than the negative control. The elderly, especially women, are prone to develop low-titered autoantibodies in the absence of clinical autoimmune disease. A panel of 11 anti-ENA antibodies including SSA(Ro), SSB(La), RNP/Sm, Jo-1, Sm, Scl-70, Chromatin, Centromere, Histone, RNA polymerase III, and dsDNA were tested. SSA(Ro), SSB(La), RNP/Sm, and Jo-1 were detected using a solid phase bio-chip immunofluorescence assay that reports qualitative and semi-quantitative results. The assessment and interpretation of the results was following the international guideline announced by the European autoimmunity standardization initiative and the International Union of Immunologic Societies/World Health Organization/Arthritis Foundation/Centers for Disease Control and Prevention autoantibody standardizing committee. The testing principles and assay process of detecting ANA and 11 anti-ENA are very similar to the procedures described in our previous work.^18^ Testing of anti-CCP3 IgG and IgA was performed at Vibrant America Clinical Laboratory (Vibrant America, LLC, Santa Clara, CA, USA) using a commercial ELISA kit (Inova Diagnostics, San Diego, CA, USA). The interpretation of the results strictly followed the protocol provided by the assay provider companies. A seropositive autoimmune subject is who has more than one of the autoantibodies tested positive. A seronegative control is whose autoantibody testing results were negative.

### Female Sex Hormone Panel

The Female Sex Hormone Panel includes estradiol, testosterone, sex hormone-binding globulin (SHBG), prolactin, progesterone, dehydroepiandrosterone sulfate (DHEA-S), luteinizing hormone (LH), parathyroid hormone, follitropin (FSH), human insulin-like growth factor 1 (HIGF1), cortisol. Serum cortisol levels were measure using the Elecsys Cortisol II immunoassay (Roche Diagnostic, USA). The Elecsys Parathyroid hormone assay (Roche Diagnostic, USA) employs a sandwich test principle in which a biotinylated monoclonal antibody reacts with the N-terminal fragment and a monoclonal antibody labeled with a ruthenium complex reacts with the C-terminal fragment. The Elecsys Estradiol III assay (Roche Diagnostic, USA) contained two monoclonal antibodies specifically directed against 17β-estradiol. The Elecsys FSH assay and LH assay (Roche Diagnostic, USA) comprised of two different monoclonal antibodies specifically directed against human FSH and human LH respectively. The Elecsys Testosterone II assay (Roche Diagnostic, USA) is employed a monoclonal antibody specifically directed against testosterone. The Elecsys SHBG assay (Roche Diagnostic, USA) employed two monoclonal antibodies specifically directed against human SHBG. The Elecsys DHEA-S (Roche Diagnostic, USA) assay comprised of a monoclonal antibody specifically directed against DHEA-S.

### Data Analysis

Clinical data from the de-identified subjects were included in a database that was processed and analyzed using Java for Windows version 1.8.45. Two-tail student T test was performed to determine whether there is significant difference between data sets and P<0.05 is considered as significant. Box plots presented the serum hormone levels in pre- and post-menopausal female seropositive autoimmune subjects and matched controls. The box represented the interquartile range (IQR), the line within the box was the median value and the whiskers show values within 1.5 IQR of the adjacent quartile. Outliers were plotted.

## RESULTS

### Patient Clinical Characteristics

15319 female subjects who ordered ANA, ENA, anti-CCP3 and the female hormone panel between December 2015 to May 2019 were included in this retrospective study. Table 1 shows the demographics of the positive subjects and negative controls in this study. Given the expected differences in reproductive and hormonal factors across the female lifespan, we explored the distribution of all markers stratified by age (12–19 years, 20–50 years, and 51 years and over; Table 1). Among the 15319 females in this cohort, 554 (3.6%) are 12 to 19 years old, 8359 (54.6%) are 20 to 50 years old, 6404 (41.8%) are more than 51 years old. Among all subjects, 4717 (30.8%) subjects have at least one autoantibody, 7506 (49.0%) subjects have at least one hormone at abnormal range, and 2415 (15.8%) subjects carry both autoantibody and abnormal hormone.

**Table 1.** Demographics of the studied subjects.

|  | All subjects | 12~19-year-old Group | 20~50-year-old Group | >50-year-old Group |
| --- | --- | --- | --- | --- |
| No. of subjects | 15319 | 554 | 8359 | 6404 |
| Average age (±SD) | 47±15 | 16±2 | 37±8 | 62±8 |
| No. of subjects with autoantibody | 4717 | 157 | 2275 | 2285 |
| No. of subjects with abnormal hormone | 7506 | 303 | 4083 | 3120 |
| No. of subjects with autoantibody and abnormal hormone | 2415 | 88 | 1124 | 1203 |

We limited subsequent analyses to women over age 20 and stratified by menopause status, due to the fewer number of subjects in the young group. Furthermore, comparing the females at their child-bearing period (20-50 years old) and menopausal period (>50 years old) yields more significant result because those are the age groups that hormones are clearly at two different stages. In the 20∼50-year-old group, 27.2% subjects have at least one autoantibody, which is significantly lower (p<0.05) than that (35.7%) of >50-year-old group. Surprisingly, the prevalence of hormonal imbalance is very close in the two age groups (48.8% and 48.7%, respectively). Occurrence of both autoantibodies and abnormal hormones is slightly more frequent in the >50-year-old group (18.8%) than that in the 20∼50-year-old group (13.4%).

### ANA, ENA, and CCP3 Autoantibodies

Self-reactive antibodies to ubiquitous cellular components are considered to be a hallmark of systemic autoimmune diseases, and ANA are the most common assessed in clinical practice. Although ANA may precede the development of systemic autoimmune disease in some cases, ANA are also frequently detected in healthy individuals and are generally considered to be non-specific markers of autoimmunity in the absence of other clinical and laboratory features of autoimmune disease. In the 1999–2004 National Health and Nutrition Examination Survey for a cohort of 351 women, the ANA test was positive with an overall weighted prevalence of 17.6%, increasing from 12.4% for ages 12∼19 to 17.4% and 19.6% for ages 20∼49 and >50, respectively.^19^ In the current cohort, the frequency of ANA positivity is 18.5% in the 20∼50-year-old group and 18.9% in the >50-year-old group (p=0.3), as shown in Figure 1.

**Figure 1.**
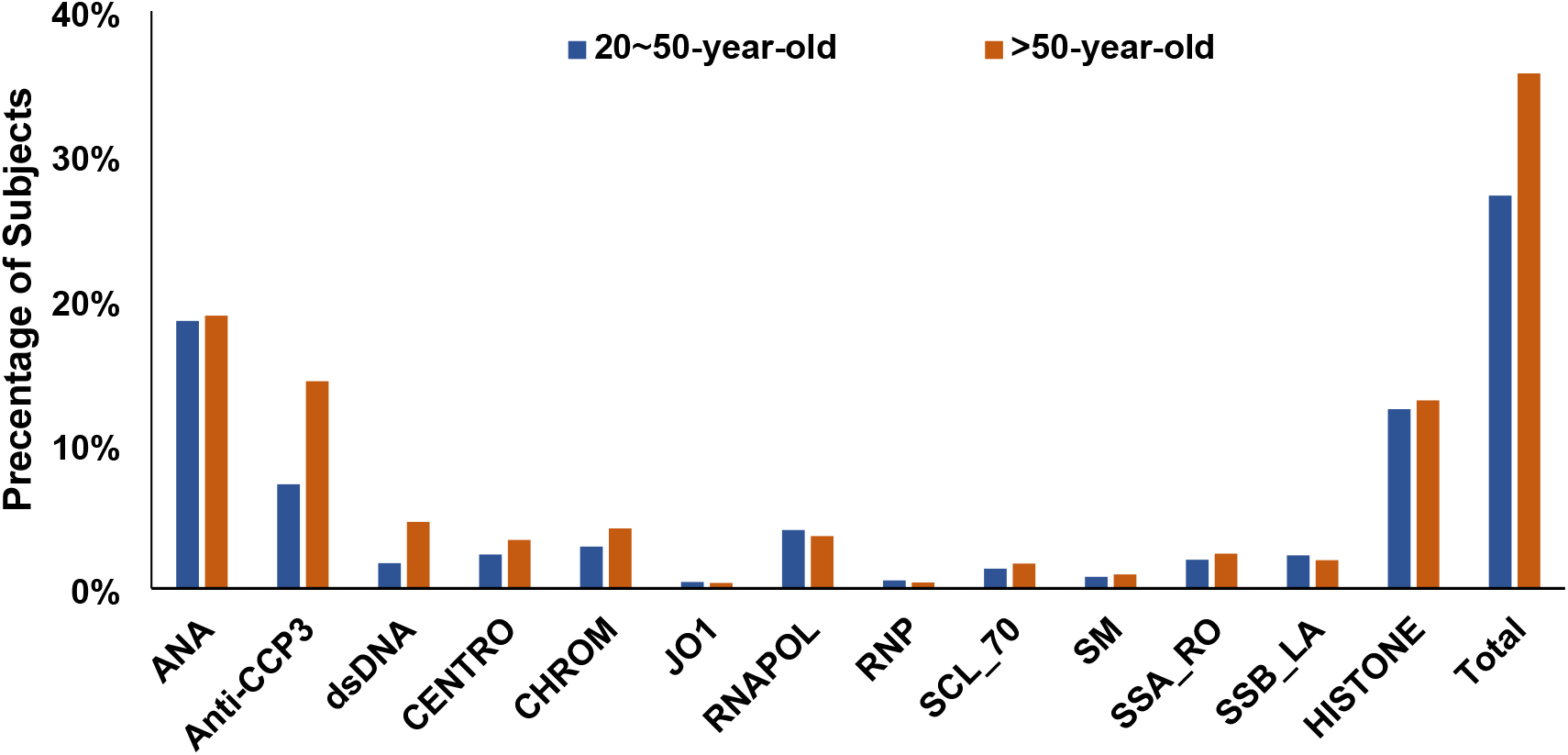
Frequency of autoantibodies’ positivity in two female age groups divided by menopause status.

Detection of anti-ENA antibodies, as a second-tier test, is usually followed by a positive ANA test to identify and distinguish between different autoimmune diseases, especially in connective tissue disorders. However, the subjects in this cohort ordered a multiplex panel in which the anti-ENAs were tested simultaneously with ANA.^18,20^ A total of 11 anti-ENAs were included in this multiplex panel and no significant difference was observed between two age groups among them. dsDNA, a critical serological marker for SLE, was included as well. Positivity in dsDNA increased from 1.7% in the 20∼50-year-old females to 4.6% in the >50-year-old females. Anti-CCP3 is a highly specific marker in Rheumatoid Arthritis patients. In this cohort, the level of anti-CCP3 in the post-menopausal group (14.4%) is significantly higher (p<0.001) than that in the child-bearing group (7.2%). If assuming any of the above-mentioned autoantibody’ positivity indicating risk in autoimmune diseases, the 20∼50-year-old group is at 27.2%, which is lower than the 35.7% in the >50-year-old group (Figure 1).

The presence of multiple autoantibodies in the two groups has also been investigated (Figure 2). The frequency of one autoantibody being positive is 20.1% in the 20∼50-year-old group and 24.6% in the >50-year-old group. There is a higher chance for female to have more than 2 autoantibodies’ positivity after the menopause age as shown in this cohort. There are 8% and 2.3% of post-menopausal females had two and three autoantibodies that are greater than the reference range, compared with 4.6% and 1.8% in the birth-bearing females.

**Figure 2.**
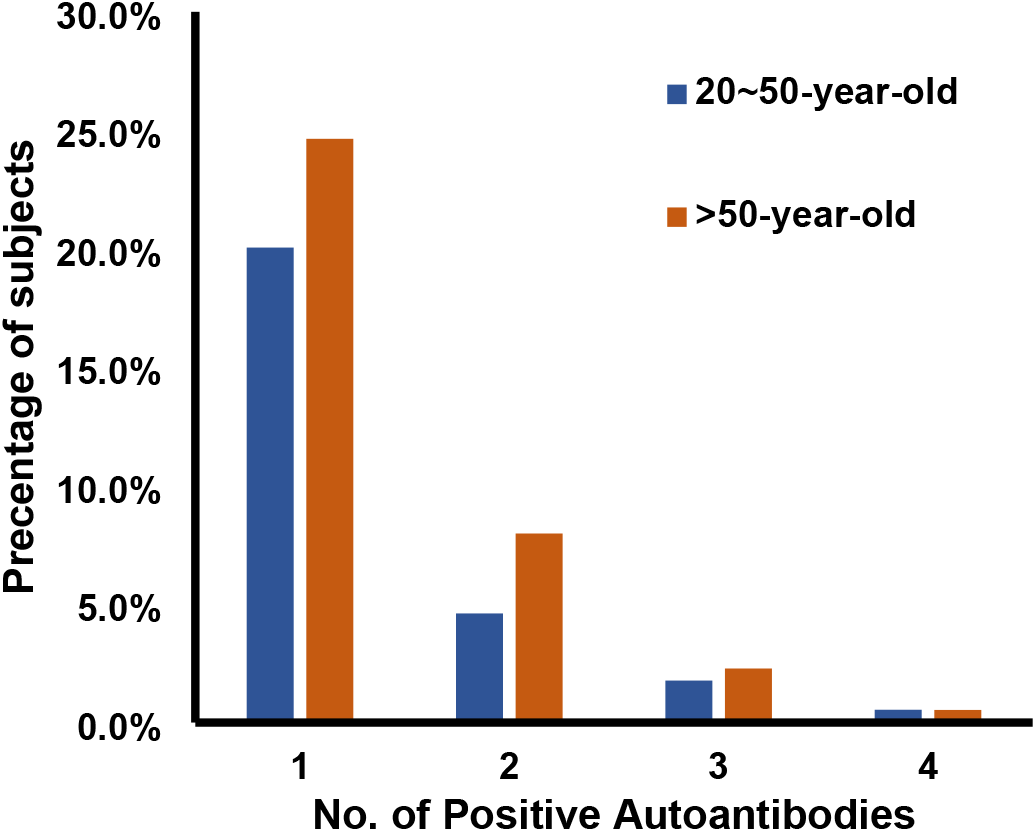
The frequency of multiple autoantibodies’ presence in two age groups.

### Estrogen and Autoantibodies

As shown in Figure 3, equivalent results were observed in the present cohort that no statistically different levels of estradiol were observed among seropositive (ANA+ or ENA+) SLE subjects and their controls. In addition, estradiol has been reported to have immune-protective effects in other autoimmune diseases such as multiple sclerosis (MS)^21^ and rheumatoid arthritis (RA).^22^ We, however, observed opposite tendency in the seropositive (CCP3+) RA subjects in both age groups. The median numbers of estradiol levels in CCP3+ subjects and controls are 72.5 pg/mL compared to 49.3 pg/mL in the 20∼50-year-old group and 17.2 pg/mL compared to 11.3 pg/mL in the >50-year-old group (Figure 3, P<0.05).

**Figure 3.**
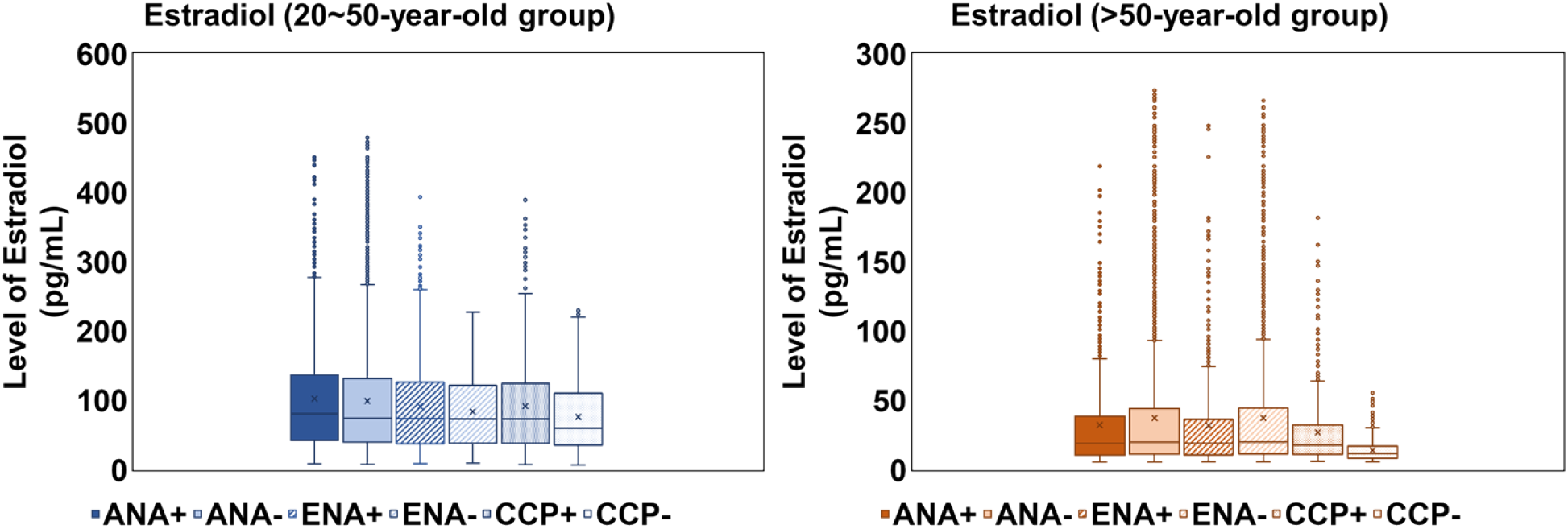
Levels of serum estradiol concentrations in pre- or post-menopausal female seropositive autoimmune subjects and matched controls.

#### Other Sex Hormones and Autoantibodies

While estrogen in general has immunostimulatory effects, the roles of other sex hormones were also widely studied in immune responses and autoimmune diseases with an emphasis on SLE. Here we consider the subject is autoimmune seropositive (AI+) if any of the ANA, anti-ENA, or anti-CCP3 markers is positive while the subject needs to be negative for all these markers to be considered as seronegative (AI-). Other than estrogen, 9 sex hormones investigated in this study include testosterone, SHBG, prolactin, progesterone, DHEA-S, LH, parathyroid hormone, FSH, HIGF1.

The serum testosterone levels were investigated in this study for AI+ and AI-female subjects across two age groups, as shown in Figure 4. In the 20∼50-year-old group, the median numbers of testosterone levels are very close among AI+ (20.1 ng/dL) and AI-subjects (21.2 ng/dL). Similarly, the >50-year-old group also has similar median numbers of testosterone levels in both subjects (AI+: 15.3 ng/dL, AI-: 15.9 ng/dL). However, no significant difference was observed between the seropositive subjects and negative controls in either of the age groups. A related hormone is SHBG, which controls the amount of testosterone that the body tissues can use. Compared to men, women naturally have higher levels of SHBG. In the current cohort shown in Figure 4, the median numbers of SHBG level in the four subgroups were also very similar (80.3, 85.6, 78.1, and 79.8 nmol/L, P>0.05). Interestingly, after menopause the serum testosterone concentration is more distributed while the SHBG level is more constrained.

**Figure 4.**
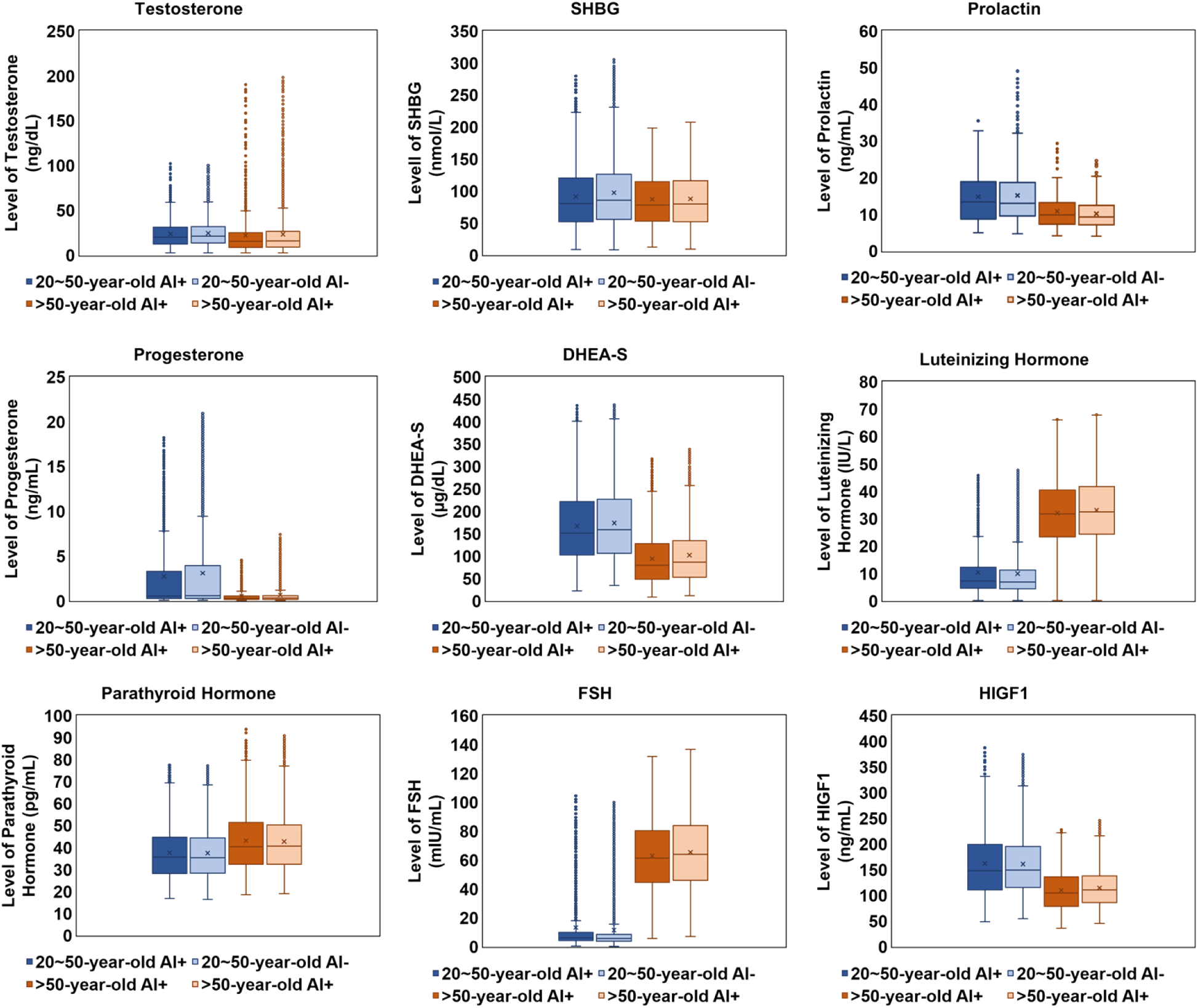
Levels of serum testosterone, SHBG, prolactin, progesterone, DHEA-S, LH, parathyroid hormone, FSH, HIGF1 in pre- or post-menopause female seropositive autoimmune subjects and matched controls.

Prolactin, as a polypeptide pituitary sex hormone, has a bioactive function acting as both a hormone and a cytokine.^23^ Prolactin has many immunoregulatory functions, mainly as an inhibitor for the negative selection of autoreactive B lymphocytes. Hyperprolactinemia has been described in relation to the pathogenesis and activity of several autoimmune disorder.^24^ Several studies have examined the serum prolactin concentrations in adult lupus patients compared with controls and disparate results were reported.^25,26^ In the current study as shown in Figure 4, the levels of prolactin were very close among the seropositive subjects and negative controls with a significant difference between two age groups (median number 13.4 and 13.0 ng/mL vs. 9.8 and 9.3 ng/mL, P>0.05).

Progesterone is another important endogenous steroid hormone that is involved in the menstrual cycle, pregnancy, and embryogenesis for women. Progesterone is an upstream precursor of testosterone and estradiol, which complements some effects of estrogen and also works with testosterone. Progesterone receptors have been found to be present in lymphoid organs and cells of the innate and adaptive immune systems.^27^ In fact, progesterone is directly involved in a certain type of autoimmune disease called Autoimmune Progesterone Dermatitis (APD), in which a recurrent skin rash generally appears during the second half of the cycle when levels of progesterone begin to rise and subsides shortly after menstruation.^28^ Previous study revealed a lower level of progesterone in SLE patients.^29^ However, in our examination (Figure 4), the younger group has higher level of progesterone (median number 0.54 ng/mL in AI+ and 0.59 ng/mL in AI-), compared to that of the senior group (median number 0.29 ng/mL in AI+ and 0.30 ng/mL in AI-) while there is no significant difference between the seropositive subjects and seronegative controls in respective groups (P>0.05).

Dehydroepiandrosterone sulfate (DHEA-S) is an adrenal androgen produced in the adrenal gland which helps produce progesterone, testosterone, and estradiol. Several studies have reported the serum levels of DHEA or DHEA-S to be lower in patients with inflammatory disease including lupus, and these levels seem to inversely correlate with disease activity.^30^ In the current cohort, we observed the same trend in both age groups that the AI+ subjects have relatively lower levels of DHEA-S compared with AI-subjects. In the 20∼50-year-old group, the AI+ subjects’ median number of DHEA-S level is 151.0 μg/dL compared with 159.1 μg/dL of the AI-subjects (P<0.05). As the age increased to more than 50 years old, the AI+ subjects’ median number of DHEA-S level decreased to 79.6 μg/dL compared with 86.5 μg/dL for the AI-subjects (P<0.05).

An acute rise of luteinizing hormone (LH) in females triggers ovulation and development of the corpus luteum. Studies of Ovarian failure and autoimmunity have detected autoantibodies directed against the unoccupied LH/human chorionic gonadotropin receptor.^31^ As observed in Figure 4, the senior groups’ median numbers of LH levels (31.7 IU/L for AI+, 32.5 IU/L for AI-) are 4 times of those in the younger age (7.2 IU/L for AI+, 6.9 IU/L for AI-).

Parathyroid hormone, also called parathormone or parathyrin, is a hormone secreted by the parathyroid glands that regulates the serum calcium through its effects on bone, kidney, and intestine. Autoimmune hypoparathyroidism develops when the body’s own immune system mistakenly attacks parathyroid tissue and leads to the loss of the secretion of parathyroid hormone. In the presented cohort, the level of parathyroid hormone is 40.3 and 40.5 pg/mL in seropositive and negative subjects respectively for ages >50, increasing from 35.6 and 35.3 pg/mL for age 20∼50 group.

Follicles produce estrogen and progesterone in the ovaries and help maintain the menstrual cycles in women. Follicle-stimulating hormone (FSH) is an important part of the reproductive system and it’s responsible for the growth of ovarian follicles. Reproductive autoimmune failure has found to be associated with overall activation of immune system or with immune system reactions specifically directed against ovarian antigens. Majority of the antiovarian autoantibodies are directed against β-subunit of follicle stimulating hormone (anti-FSH).^32^ As observed in Figure 4, after the menopause, the median number of FSH levels immediately increased from 6.1 and 5.78 mIU/mL to 61.2 and 63.8 mIU/mL for AI+ and AI-subjects, respectively.

Human insulin-like growth factor 1 (HIGF1), also called somatomedin C, is a hormone similar in molecular structure to insulin which plays an important role in childhood growth and has anabolic effects in adults. Recent data highlight a significant interaction of IGFs and the components of the immune system, especially B and T cells. Accumulating data suggest an important role of IGFs in autoimmune diseases, including SLE, RA, systemic sclerosis and Sjogren’s syndrome.^33^ In the current cohort, there is not significant difference between AI+ and AI-subjects’ HIGF1 level among the 20∼50-year-old group (147.8 and 149.0 ng/mL); however, the AI-subjects have a lower level of HIGF1 (median number 104.5 ng/mL) than in AI+ subjects (median number 110.8 ng/mL).

### Cortisol and Autoantibodies

Other than sex hormones, cortisol, as a steroid hormone, has also been shown in previous studies to be affect in an autoimmune disease, namely Primary Adrenal Insufficiency (Addison’s disease). Addison’s disease is a rare disorder characterized by inadequate production of the cortisol and aldosterone by the two outer layers of cells of the adrenal glands (adrenal cortex).^34^ Addison’s disease occurs when the body’s immune system mistakenly attacks the adrenal glands causing slowly progressive damage to the adrenal cortex. In the current study, no significant difference was observed between AI+ and AI-subjects across both age groups while the younger group has a much-scattered distribution as shown in Figure 5.

**Figure 5.**
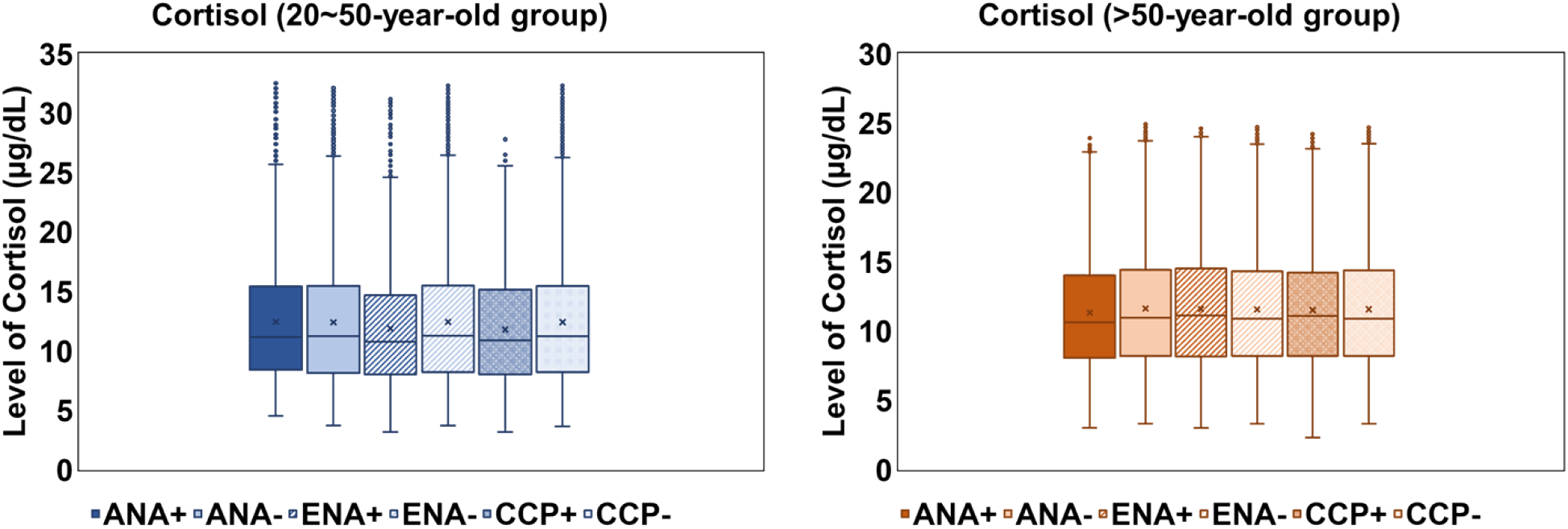
Levels of serum cortisol in pre- or post-menopausal female seropositive autoimmune subjects and matched controls.

## DISCUSSION

The strongest risk factor for development of some of autoimmune diseases (e.g., SLE, RA, MS, etc) appears to be female sex. Take SLE as an example, the female-to-male sex ratio is 9:1 during the peak reproductive years of females and the ratio gradually decline after menopause, while the age of diseases onset for males is more evenly distributed. The correlations between the autoimmune disease activity and sex hormone concentrations have been discussed for decades but relatively few patients and controls have been examined. A clear understanding of relationships between serum sex hormonal concentrations and development of autoimmune diseases remains elusive due to variability and non-homogeneity of the currently available data. However, majority of these comparison studies did not observe statistically different results for the sex hormones, as well as in the presented study. In fact, the serum sex hormone concentrations are typically within the physiologic ranges for patients with autoimmunity, although the serum levels of certain hormones were also found to be statistically higher or lower than those in matched controls. In this study, we, as a clinical laboratory, explored the levels of serum sex hormones within a large cohort of female subjects with seropositive autoimmunity or matched negative controls. The pre- or post-menopausal status is a further possible factor influencing the rate of autoimmune diseases; therefore, the epidemiologic data in the current study was broken down into two ages and female-specific groups before making comparisons.^35^

While serum estrogen levels have not been found to be significantly different in women with SLE as observed in this study and several investigations, increased estrogen metabolism has been observed.^36^ Higher levels of more feminizing estrone metabolites were observed in SLE patients and their first-degree relatives implying that more potent metabolites may induce more potently epigenetic changes via the estrone receptors. By counteracting the pathways of estrogen, androgens and progesterone are widely recognized as natural immune suppressors. Progesterone has been shown to reduce T cell proliferation and impact T cell-dependent antibody responses,^37^ while low testosterone levels are correlated with higher B cells and antibody responses.^38^ In the current study, however, no significantly different levels of testosterone and progesterone were observed in seropositive (AI+) subjects comparing with the controls within respective age groups. Less frequently investigated sex hormones including SHBG, Prolactin, Parathyroid Hormone, Luteinizing Hormone, DHEA-S, FSH, HIGF1 were also explored in this study. The limitation of the current study is that several physiological, pathological, and therapeutic conditions may change the serum sex hormones milieu and/or peripheral conversion rate, including the menstrual cycle, pregnancy, postpartum period, menopause, being elderly, chronic stress, altered circadian rhythms, inflammatory cytokines, and use of corticosteroids, oral contraceptives, and steroid hormonal replacements, inducing altered androgen/estrogen ratios and related effects.

In conclusion, the comparison studies in the presented study involving a large cohort showed no statistically different levels of sex hormones in seropositive autoimmune subjects and matched controls except for DHEA-s, which is in accordance with previous studies on small cohorts. Our study shows that even though sex hormones might be an important factor of autoimmune disease based on epidemiologic evidence, the correlation seems to be indirect and pathogenesis pathway needs systemic investigation. Evidence remains to be explored in the complex interactions of hormones, genetic factors, and environmental factors in individuals with deregulation of the immune response including autoimmune diseases.

## Data Availability

All relevant data are within the paper. Any additional data will be available upon request from Vibrant Sciences LLC by sending an email to.

## DECLARATIONS

### Authors’ contributions

VJ, HK, JJR designed the study; YY, HK wrote the manuscript; KK, TW collected and tested the samples; YY, QS collected the clinical data; YY analyzed the data; KB developed the Vibrant TSP Software. All the authors read and approved the final manuscript.

## Acknowledgement

We acknowledge Vibrant America LLC for supporting this research.

## Competing interests

YY, QS are employees of Vibrant America LLC. KK, VJ, TW, KB, HK, JJR, are employees of Vibrant Sciences LLC.

## Availability of data and material

The data used to support the findings of this study can be acquired from Vibrant America LLC.

## Ethics approval and consent to participate

IRB exemption (work order #1-1098539-1) was determined by the Western Institutional Review Board (WIRB) for Vibrant America Biorepository to use de-linked and de-identified remnant human specimen and medical data for research purposes.

## Consent for publication

NA

## Funding

Vibrant America LLC.

## Notes

### Competing Interest Statement

The authors have read the journal's policy and the authors of this manuscript have the following competing interests: YY and QS are paid employees of Vibrant America LLC. KK, VJ, TW, KB, HK, JJR, are paid employees of Vibrant Sciences LLC. Vibrant Sciences or Vibrant America could benefit from increased testing based on the results. There are no patents, products in development, or marketed products to declare. This does not alter our adherence to the journal policies on sharing data and materials.

### Author Declarations

Western Institutional Review Board (WIRB)

